# Endogenous Retroelement Expression in the Gut Microenvironment of People Living with HIV-1

**DOI:** 10.1101/2023.11.06.23298166

**Authors:** Nicholas Dopkins, Tongyi Fei, Stephanie Michael, Nicholas Liotta, Kejun Guo, Kaylee L. Mickens, Brad S. Barrett, Matthew L. Bendall, Stephanie M. Dillon, Cara C. Wilson, Mario L. Santiago, Douglas F. Nixon

## Abstract

**Background:** Endogenous retroelements (EREs), including human endogenous retroviruses (HERVs) and long interspersed nuclear elements (LINEs), comprise almost half of the human genome. HIV-1 infects a target cell already possessing ancient retroviral genetic material, and exogenous HIV-1 infection modulates the expression of cell associated EREs. Following initial HIV-1 infection, there is a rapid destruction of CD4+ cells in the gut associated lymphoid tissue (GALT). Our previous studies of the interferome in the gut suggest potential mechanisms regarding how IFNb may drive HIV-1 gut pathogenesis. As ERE activity is suggested to partake in type 1 immune responses and is incredibly sensitive to viral infections, we sought to elucidate underlying interactions between ERE expression and GALT dynamics in PLWH.

**Methods:** ERE expression profiles from bulk RNA sequencing of colon biopsies and PBMC were compared between a cohort of PLWH prior to or within 7 days of initiating antiretroviral therapy (ART) (n=19) and uninfected controls (n=13). Individual EREs were then compared with the profiling of uninfected gut CD4+ T cells activated with type 1 interferons (IFN-Is) (n=3) to elucidate potential mechanisms for their induction in PLWH.

**Findings:** 59 EREs were differentially expressed in the colon of PLWH when compared to uninfected controls (Wald’s Test with Benjamin-Hochberg correction: padj < 0.05 and FC ≤ −1 or ≥ 1). Of these 59, 12 EREs were downregulated in PLWH and 47 were upregulated. Colonic expression of the ERE loci LTR19_12p13.31 and L1FLnI_1q23.1s showed significant correlations with CD8+ T Cells and dendritic cell subset frequencies in the GI tract (Spearman’s Correlation: p value < 0.05). Furthermore L1FLnI_1q23.1s showed a significant upregulation in the blood of PLWH when compared to uninfected controls (T test: p <0.05) suggesting a common mechanism of differential ERE expression in PBMC and GALT.

**Interpretation:** ERE activity has been largely understudied in genomic characterizations of human pathologies. We show that the activity of certain EREs in the GI tract of PLWH is deregulated, supporting our hypotheses that their underlying activity could function as (bio)markers and potential mediators of pathogenesis in HIV-1 reservoirs.

**Funding:** NCI CA260691 (DFN) and NIAID UM1AI164559 (DFN).

## Introduction

Human immunodeficiency virus type I (HIV-1) is a retrovirus that infects immune cells by binding to receptors on the cell surface^1^. HIV-1 infects host cells by integrating a copy of the viral genome into the host genome, resulting in proviruses that then hijack the host cell’s replicative machinery to produce the building blocks of infectious particles^1^. The HIV-1 lifecycle may rapidly deplete host CD4+ T cell populations and disrupt global immune dynamics^2^. Disruptions to the host immune system predispose people living with HIV-1 (PLWH) to life-threatening comorbidities^3^. Since 1996, life expectancy and quality of life for PLWH have risen due to the advent of antiretroviral therapies (ARTs) that pharmacologically inhibit key processes of the retroviral life cycle^4^. Unfortunately, ART merely suppresses retroviral processes, emphasizing the need for cure strategies that rid individuals of latent proviral reservoirs which may resume infectious particle formation after a lapse in treamtent^5,6^. One such reservoir site is the gastrointestinal (GI) associated lymphoid tissue (GALT)^7,8^, which is particularly rich in CD4+ T cells^9^. HIV-1 induced immunopathologies in the GALT are associated with severe comorbidities in PLWH^10,11^. Many of these pathologies could be sustained by the accumulation of plasmacytoid dendritic cells (pDCs) that are trafficked to the GI tract during chronic HIV-1 infection^12^. Models of Simian Immunodeficiency Virus (SIV), a retrovirus closely related to HIV-1, demonstrate that pDCs traffic from the circulation, where they are depleted, to the GALT, where their overabundance contributes to immunopathologies that allow the site to become the primary source of viral replication^13,14^. pDCs reflect a unique subset of DCs whose primary function is not antigen presentation but is instead immunomodulation through the release of cytokines^15^. In the acute stages of HIV-1 infection, pDCs produce type 1 interferons (IFN-Is) as one of the first lines of defense against the virus^16^, and are therefore protective. However, the sustainment of their IFN-I production during the chronic stages of infection exhausts and deregulates antiviral immunity, negatively impacting the health of PLWH^12,16–18^.

To better characterize how immunopathologies in PLWH develop, we investigated how endogenous retroelement (ERE) expression may underlie the development of HIV-1 induced immune deregulation. EREs are highly abundant sequences estimated to make up roughly ∼40% of the human genome^19^. In humans, EREs include long interspersed nuclear elements (LINEs), short interspersed nuclear elements (SINEs), and human endogenous retroviruses (HERVs)^20^. Previous studies have demonstrated that ERE activity is highly susceptible to change during infection with HIV-1^21–31^. While IFN-I signaling may activate EREs proximal to interferon stimulated genes (ISGs), other EREs, such as HERV elements, can be directly trans-activated by HIV-1 cofactors due to high sequence homology between endogenous and exogenous retroviruses^21,22,24,32,33^. This homology can be so substantial that certain HERV-K18 and HERV-W products can be incorporated in the assembly of HIV-1 particles^34–38^. ERE activity also functions as a major regulator of transcriptional networks that have been co-opted in organismal physiology^39,40^. In immunity, examples of ERE activity cooption suggest their intimate association with the host mounted responses^41,42^. Recurrent observations continually reinforce hypotheses that EREs likely participate in human health and disease through poorly understood mechanisms^42–46^. The activity of multiple HERVs^21–28^ and LINEs^29,30^ is altered during HIV-1 infection, which then impacts host gene expression^24^, packaging of HIV-1 particles^34–38^, and immunity^25,47^. For the purpose of further defining HIV-1 mediated gut pathogenesis, we sought to characterize how ERE activity at the RNA level is modulated in the gut microenvironment of PLWH. Since EREs have been largely overlooked in genomic approaches to classify underlying components of human pathologies, we hypothesize that their consideration would identify novel markers of HIV-1 pathogenesis in the gut.

ERE expression at the RNA level was quantified in colon biopsies collected from a cohort of viremic individuals prior to or at the initiation of ART^17^ to identify those best correlating with viral load. We next overlaid these findings with IFN-I stimulated gut CD4+ T cells to estimate if the differential expression of EREs in the gut of PLWH is driven by the HIV-1 lifecycle or by innate immunopathologies. Collectively, our analyses demonstrate that a subset of ERE loci are impacted in PLWH. The bulk of PLWH-upregulated EREs in the gut are not upregulated by IFN-I stimulation of gut CD4+ T cells, suggesting their activation by other aspects of the HIV-1 life cycle or the host response. Two identified EREs, LTR19_12p13.31 and L1FLnI_1q23.1s, further correlated with myeloid and lymphoid immune cell abundances. Collectively, incorporation of the GI-specific expression of LTR19_12p13.31 and L1FLnI_1q23.1s can better define HIV-1 driven immunopathologies of the GALT, and the expression of these elements may possess underlying roles in pathogenesis.

## Methods

### Locus-specific quantification of ERE expression at the RNA level

RNA sequencing FASTQ files of colon pinch biopsies and interferome arrays of uninfected CD4+ T cells were obtained from a previously published study^17^ under the Bioproject accession numbers “PRJNA558500” and “PRJNA558974”, respectively. RNA sequencing FASTQ files performed on the PBMC fraction from the same participants are available under the Bioproject accession number “PRJNA659373”. FASTQ files were aligned to the human genome build 38 (hg38) using STAR (v2.7.9.a)^48^. STAR alignment was performed using the parameters “--outSAMstrandField intronMotif --outFilterMultimapNmax 200 --winAnchorMultimapNmax 200” for the purpose of capturing multimapping ERE reads. We then used Telescope (v1.0.3)^49^, which utilizes an expectation-maximization algorithm to improve upon the definition of ubiquitously mapping ERE transcripts to a custom annotation, for locus-specific approximation of ERE expression. The Telescope assign module was performed using the parameters “--theta_prior 200000 -- max_iter 1000” to align reads to a custom annotation of EREs accessible at https://github.com/mlbendall/telescope_annotation_db. Metadata for the ERE annotation predicting the intronic, exonic, and intergenic status of individual loci in the Telescope annotation relative to coding gene annotations can be found at https://github.com/liniguez/Telescope_MetaAnnotations. Original code produced for analysis can be accessed at https://github.com/NicholasDopkins/JuneHIVMucosa.

### Differential expression analysis

Lowly abundant EREs were filtered from downstream analysis by ensuring that all EREs possessed at least 2 reads within 10% of the total samples. Differential expression analysis was performed between the ERE expression profiles of uninfected control and PLWH colon biopsies using standard DESEQ2 (v1.30.1)^50^ parameters. Differential expression analysis was performed between the ERE expression profiles of IFN-I stimulated gut CD4+ T cells using DESEQ2 with the parameters “parallel = T” and “betaPrior = T”. Results were extracted as DESEQ objects, with a numbered contrast of each group compared against all others. Differentially expressed EREs were visualized with the packages pheatmap (v1.0.12), ggVennDiagram (v1.2.2)^51^ and EnhancedVolcano (v1.8.0). Individual EREs were visualized with a custom function that overlays geom_jitter and geom_boxplot functions provided by ggplot2 v(3.3.5)^52^ to produce a Tukey box and whisker plot that displays individual replicates. Clinical parameters of immunopathologies were provided by previously published metadata^17,18,53,54^ and a table summary is available at https://github.com/NicholasDopkins/JuneHIVMucosa/blob/main/Metadata.csv. Principal coordinate analyses (PCA) were performed on variance stabilizing transformed DESEQ outputs in order to visualize if untested confounding factors explained observations pertaining to differential expression analyses. PCA generation was performed using PCAtools (v2.2.0) with the variable parameter set to “removeVar = 0.1”.

### Integrative Genomics Viewer

National Center for Biotechnology Information (NCBI) Reference Sequence Database (Refseq)^55^, ENCyclopedia Of DNA Elements^56^ (Gencode), and Telescope^49^ annotations of the human genome were loaded into Integrative Genomics Viewer^57^ (IGV) for alignment of StringTie^58^ reconstructed transcriptomes from uninfected controls and PLWH samples.

### Statistics

Standard DESEQ2 statistics of Wald’s Test with Benjamin-Hochberg correction of p values were utilized for the identification of significantly deregulated EREs between uninfected controls and PLWH samples. EREs a log2 fold change of >1 or <-1 and an adjusted p value (padj) of <0.05 were deemed significantly differentially expressed. Statistical values for correlations between clinical parameters and ERE expression were conducted using Pearson’s correlation coefficient values provided by ggplot2. To elucidate values of significance regarding of ERE expression between experimental groups, T tests were used to gather significance values when stated. For significance of post hoc tests the following scale is used to indicate p values when applicable: ns, p > 0.05; *, p < 0.05; **, p < 0.01; ***, p < 0.005

### Clinical cohort

ERE expression analyses were conducted using FASTQ files from previously generated RNAseq data collected from colonic tissue and PBMC fractions collected from 19 PLWH and 13 age- and sex-matched controls, and associations with features of HIV-1 pathogenesis were determined using archived datasets of colon and systemic immunological and virological parameters^17,18,53,54^. On average, PLWH had been infected with HIV-1 (defined by the first HIV-1 seropositive test) for 5.25±1.1 (mean±SEM) years. PLWH were ART-naive or were not on ART for more than 7 days in the preceding 6 months, and had CD4 T cell counts >200 cells/μl within 3 months of clinical visit. Exclusion criteria for both cohorts are extensively detailed elsewhere^53^. In addition to blood samples, study participants underwent a flexible sigmoidoscopy with multiple colon pinch biopsies obtained. Clinical characteristics for the study participants included in this current study are detailed in Supplemental Table 1.

### Immune system characterization of PLWH and uninfected controls

Methods used to generate the archived datasets have been extensively detailed^17,18,53,54,59,60^. In brief, indicators of systemic inflammation were assessed with plasma IL-6 concentrations by ELISA, and microbial translocation was measured by plasma LPS concentrations with the Limulus Amebocyte Lysate assay^53^. Frequencies of colon mDCs, pDCs, and T cells were determined using multi-color flow cytometry^53,54^ and expressed as an absolute number per gram of tissue based on the frequency within viable CD45+ cells, initial cell counts, and biopsy weights. Colon tissue HIV-1 RNA was quantified by real-time PCR and HIV-1 RNA copy numbers normalized per CD4 T cell within each biopsy calculated by the percent of all viable cells that were CD45+CD3+CD4+ as determined by flow cytometry and weight of each biopsy and reported as HIV-1 RNA per million CD4 T cell^53^.

### Ethics Statement

Analyses performed in this study were performed on deidentified and publicly available or previously archived data. The original clinical study was approved by the Colorado Multiple Institutional Review Board and all study participants voluntarily gave written, informed consent^17,18,53,54^.

## Results

### HIV-1 status influences endogenous retroelement expression in the gut microenvironment

Following preprocessing to remove lowly abundant transcripts, we quantified the expression of 13,706 EREs in colon pinch biopsies collected from PLWH in comparison to age and sex-matched HIV-1-uninfected controls (Fig. 1a). Significance in differential expression between controls and PLWH was found for 59 ERE loci (Fig. 1b and Fig. 1c). Of these 59 loci, 47 were upregulated and 12 were downregulated in PLWH. All HERVs differentially expressed were upregulated in PLWH when compared to controls. Overall, ERE activity at the RNA level displays discrete changes in the gut microenvironment in viremic individuals prior to the initiation of ART. Further consideration of the phylogenies and chromosomal origin of ERE expression profiles demonstrated no distinctive alterations in PLWH when compared to controls, suggesting no large-scale changes in ERE expression in the gut microenvironment of PLWH (Fig. S1). To infer mechanisms by which HIV-1 infection induces the differential expression of EREs in the gut, we next quantified changes in expression resulting from IFN-I stimulation of uninfected gut-derived CD4+ T cells. IFN-Is do not shift the global landscape of ERE activity at the RNA level (Fig. S2). Discrete changes in locus-specific expression demonstrate that IFN-I activation of uninfected gut CD4+ T cells share upregulated expression of 23 EREs, suggesting that they are IFN-inducible (Fig. S3 and Fig. S4). 7 of these 23 EREs were upregulated in the gut of PLWH, suggesting immune-dependent mechanisms for their in HIV-1 infection, while 35 of the 42 upregulated EREs in PLWH showed no significant induction by IFN-I stimulation (Fig. S5). Collectively, HIV-1 infection upregulates specific EREs in the gut of PLWH through both IFN-dependent and IFN-independent mechanisms.

**Figure 1.**
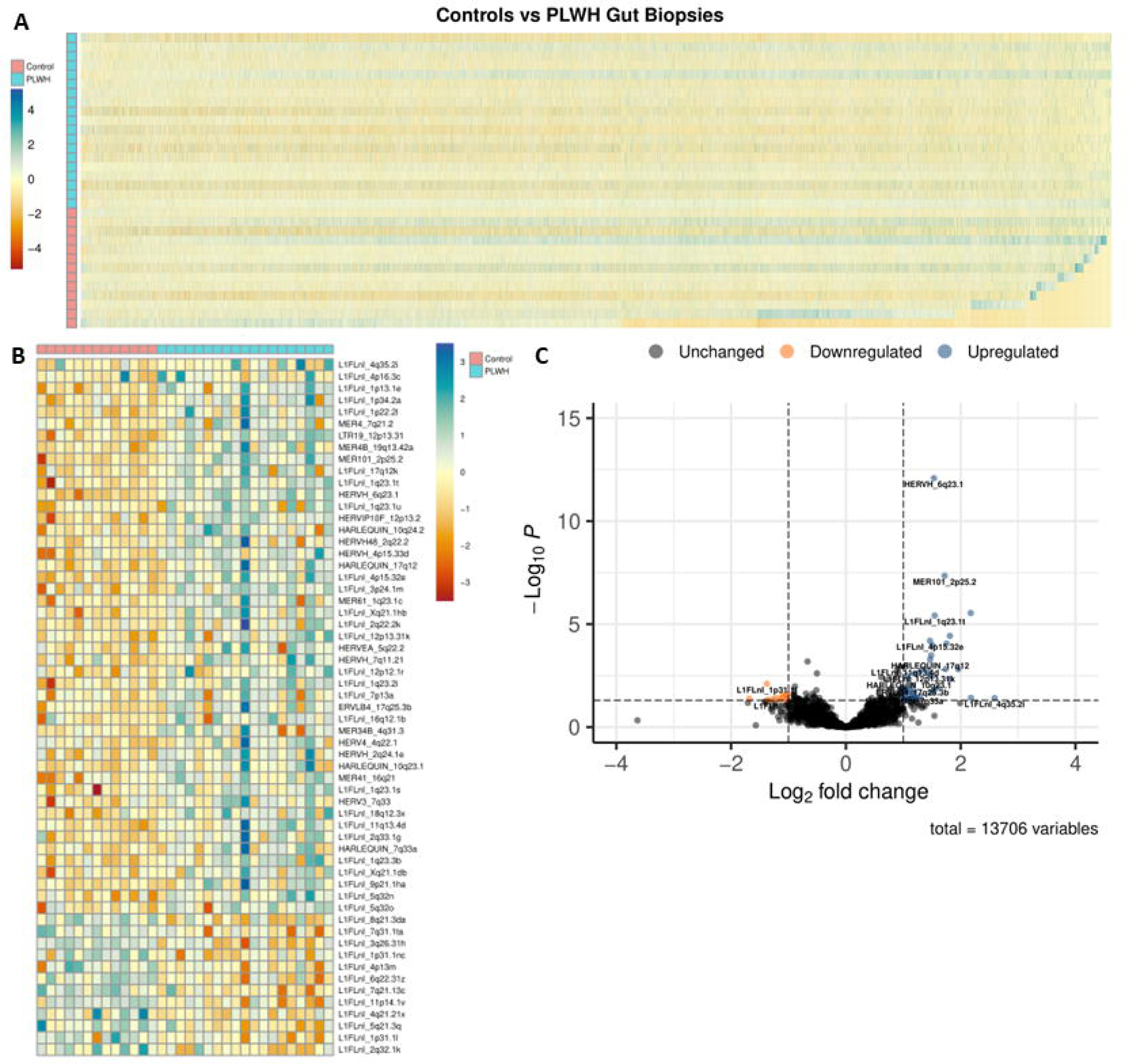
HIV-1 status influences endogenous retroelement expression in the gut microenvironment. Heatmap demonstrating the normalized per sample abundances of EREs following the preprocessing filtering of reads in uninfected controls (red) (n = 13) vs PLWH (blue) (n = 19) individuals. ERE reads were filtered to possess at least 2 reads in 10% of the total samples for quality assurance (A). Heatmap demonstrating the normalized per sample abundances of EREs differentially expressed with an adjusted p value of ≤0.05 and a LFC ≥1 or≤-1 between uninfected controls (red) (n = 13) and PLWH (blue) (n = 19) gut biopsies. All statistics were performed in DESEQ using the Wald’s Test. Adjusted p values were calculated using default parameters for a Benjamini-Hochberg correction (B). Volcano plot demonstrating the average fold change ≥1 or≤-1 and an adjusted p value of ≤0.05 and a LFC between uninfected controls (n = 13) and PLWH (n = 19) gut biopsies. All statistics were performed in DESEQ using the Wald’s Test. Adjusted p values were calculated using default parameters for a Benjamini-Hochberg correction (C).

### Correlation between gut endogenous retroelement expression and viral load in people living with HIV-1

Pearson’s correlation coefficients were calculated for all differentially expressed EREs in the gut of PLWH to identify any potential links with their expression and HIV-1 infection severity as determined by circulating HIV-1 viral load. Of the 59 differentially expressed EREs, three showed near significant (p<0.10) positive correlations between colonic expression and viral load in PLWH. L1FLnI_1q23.1s possessed a positive association with viral load in the circulation (Fig. 2a) and is upregulated in the colon of PLWH when compared to uninfected controls (Fig. 2b). In PBMCs collected from the same participants, expression of L1FLnI_1q23.1s is significantly upregulated in concurrence with gut expression of the element (Fig. 2c). Analysis of IFN-I activated gut CD4+ T cells suggests that IFN alpha 1 (IFNa1) and IFN beta (IFNb) may contribute to the upregulated expression of L1FLnI_1q23.1s, however no IFN-Is yielded significant effects (Fig. 2d). IGV demonstrates that the L1FLnI_1q23.1s locus is an ERE within the Pyrin And HIN Domain Family Member 1 (PYHIN1) gene with minor differences in the putative alignments provided by StringTie for uninfected controls and PLWH samples (Fig. 2e). L1FLnI_5q32o possessed a positive association with viral load in the circulation (Fig. 2f) and is upregulated in the colon of PLWH when compared to controls (Fig. 2g). In PBMCs collected from the same study participants, expression of L1FLnI_5q32o is significantly upregulated in concurrence with gut expression of the element (Fig. 2h). Analysis of IFN-I activated gut CD4+ T cells suggests that IFN-Is yield no significant effects on the expression of L1FLnI_5q32o (Fig. 2i). IGV demonstrates that the L1FLnI_5q32o locus is an ERE within the Janus Kinase And Microtubule Interacting Protein 2 (JAKMIP2) gene with no differences in the putative alignments provided by StringTie for PLWH and control samples (Fig. 2j). LTR19_12p13.31 possessed a positive association with viral load in the circulation (Fig. 2k) and is upregulated in the colon of PLWH when compared to controls (Fig. 2l). In PBMCs collected from the same study participants, expression of LTR19_12p13.31 shows a non-significant upregulation (Fig. 2m). Analysis of IFN-I activated gut CD4+ T cells suggests that IFN-Is yield no significant effects on the expression of LTR19_12p13.31 (Fig. 2n). IGV demonstrates that LTR19_12p13.31 locus is an ERE that overlaps the Pregnancy Zone Protein (PZP) and Killer Cell Lectin Like Receptor G1 (KLRG1) genes with a unique transcript in the putative alignments of PLWH samples provided by StringTie (Fig. 2o). Collectively, expression of the EREs L1FLnI_1q23.1s, L1FLnI_5q32o, and LTR19_12p13.31 show associations with blood HIV-1 RNA levels,, and their induction in the gut may likely be independent of IFN-I mediated pathologies.

**Figure 2.**
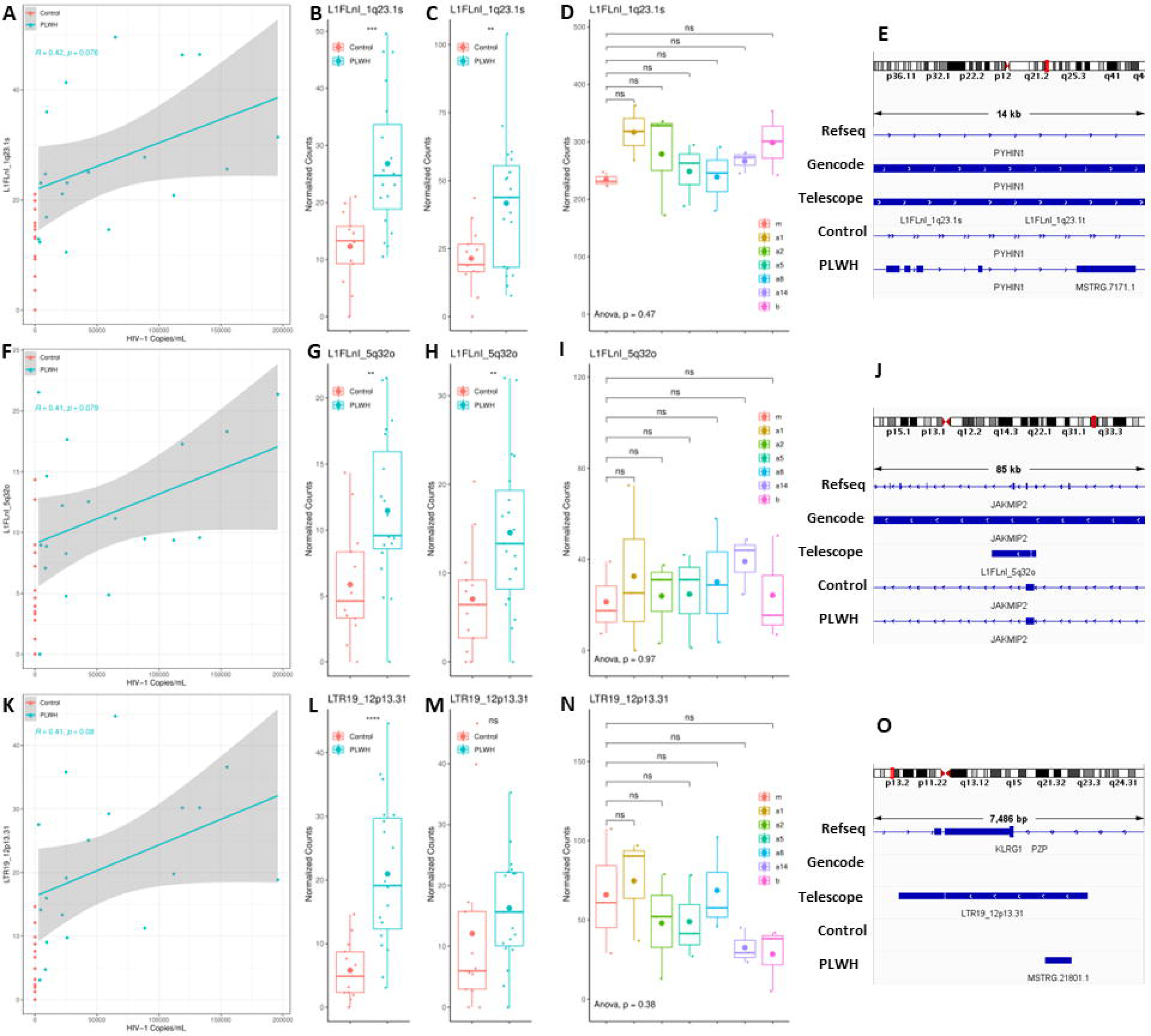
Correlation between gut endogenous retroelement expression and viral load in people living with HIV-1. Scatterplot demonstrating the viral load from serum on the X axis and normalized expression of L1FLnI_1q23.1s in colon biopsies on the Y axis. uninfected controls (red) (n = 13) vs PLWH (blue) (n = 19) (A). Normalized expression of L1FLnI_1q23.1s in colon biopsies from uninfected controls (red) (n = 13) vs PLWH (blue) (n = 19). Post hoc significance values determined with unpaired T test (B). Normalized expression of L1FLnI_1q23.1s in PBMCs from uninfected controls (red) (n = 13) vs PLWH (blue) (n = 19). Post hoc significance values determined with unpaired T test (C). Interferome array of L1FLnI_1q23.1s expression following IFN-a1 (n = 3), -a2 (n = 3), -a5 (n = 3), -a8 (n = 3), -a14 (n = 3), and -b (n = 3) treated gut CD4+ T cells when compared to mock (m) treatment (n=3). ANOVA performed for significance with post hoc significance values determined via unpaired T test (D). IGV representation of L1FLnI_1q23.1s locus provided by the Telescope annotation in the Refseq and Gencode annotations. StringTie reconstructed transcriptomes demonstrate putative transcripts present in sequencing collected from the colon of uninfected controls and PLWH individuals (E). Scatterplot demonstrating the viral load from serum on the X axis and normalized expression of L1FLnI_5q32o in colon biopsies on the Y axis. Uninfected controls (red) (n = 13) vs PLWH (blue) (n = 19) (F). Normalized expression of L1FLnI_5q32o in colon biopsies from uninfected controls (red) (n = 13) vs PLWH (blue) (n = 19). Post hoc significance values determined with unpaired T test (G). Normalized expression of L1FLnI_5q32o in PBMCs from uninfected controls (red) (n = 13) vs PLWH (blue) (n = 19). Post hoc significance values determined with unpaired T test (H). Interferome array of L1FLnI_5q32o expression following IFN-a1 (n = 3), - a2 (n = 3), -a5 (n = 3), -a8 (n = 3), -a14 (n = 3), and -b (n = 3) treated gut CD4+ T cells when compared to mock (m) treatment (n=3). ANOVA performed for significance with post hoc significance values determined via unpaired T test (I). IGV representation of L1FLnI_5q32o locus provided by the Telescope annotation in the Refseq and Gencode annotations. StringTie reconstructed transcriptomes demonstrate putative transcripts present in sequencing collected from the colon of uninfected controls and PLWH individuals (J). Scatterplot demonstrating the viral load from serum on the X axis and normalized expression of LTR19_12p13.31 in colon biopsies on the Y axis. uninfected controls (red) (n = 13) vs PLWH (blue) (n = 19) (K). Normalized expression of LTR19_12p13.31 in colon biopsies from uninfected controls (red) (n = 13) vs PLWH (blue) (n = 19). Post hoc significance values determined with unpaired T test (L). Normalized expression of LTR19_12p13.31 in PBMCs from uninfected controls (red) (n = 13) vs PLWH (blue) (n = 19). Post hoc significance values determined with unpaired T test (M). Interferome array of L1FLnI_1q23.1s expression following IFN-a1 (n = 3), -a2 (n = 3), -a5 (n = 3), -a8 (n = 3), -a14 (n = 3), and -b (n = 3) treated gut CD4+ T cells when compared to mock (m) treatment (n=3). ANOVA performed for significance with post hoc significance values determined via unpaired T test (N). IGV representation of LTR19_12p13.31 locus provided by the Telescope annotation in the Refseq and Gencode annotations. StringTie reconstructed transcriptomes demonstrate putative transcripts present in sequencing collected from the colon of uninfected controls and PLWH individuals (O).

### Correlation Between LTR19_12p13.31 and L1FLnI_1q23.1s expression and immune signatures of the gut microenvironment in people living with HIV-1

We next analyzed how expression of L1FLnI_1q23.1s, L1FLnI_5q32o and LTR19_12p13.31 correlate with cellular abundances that may be associated with pathogenesis in the gut of PLWH. There were no significant associations between the number colon CD4+ T cells (measured per gram of gut tissue)with LTR19_12p13.31 (Fig. 3a) and L1FLnI_1q23.1s expression (Fig. 3b). LTR19_12p13.31 demonstrated significant positive correlations with the number of colon CD8+ T cells (Fig. 3c) and IFN gamma (IFNy)+CD8+ T cells (Fig. 3e), whereas L1FLnI_1q23.1s demonstrated near significant positive correlations with CD8+ (Fig. 3d) and IFNy+CD8+ T cell (Fig. 3f) abundances. Further consideration into dendritic cell (DC) abundances demonstrate that LTR19_12p13.31 (Fig. 3g) and L1FLnI_1q23.1s (Fig. 3h) show near significant positive correlations with CD11c+ myeloid DCs (mDCs), while LTR19_12p13.31 (Fig. 3i) displayed a significant positive correlation and L1FLnI_1q23.1s (Fig. 3j) displayed a near significant positive association with plasmacytoid DCs (pDCs). Amongst mDCs, LTR19_12p13.31 (Fig. 3k) and L1FLnI_1q23.1s (Fig. 3l) show significant positive correlations with the CD1c-mDCs, but not with CD1c+ mDCs (Fig. S6), suggesting potential perturbations related to specific mDC subsets^54^. Expression of the ERE L1FLnI_5q32o did not demonstrate positive or negative correlations with CD4+ T cells, CD8+ T cells, IFNy+CD8+ T cells, CD11c+ mDCs, pDCs, or CD1c-mDCs in PLWH (Fig. S7). In analyses of frequencies of CD4 T helper (Th) cell subsets, L1FLnI_1q23.1s significantly positively correlated with IL22+IFNy-IL17-CD4+ T cells (i.e. Th22) and IFNy-IL17+ CD4+ T cells (i.e Th17), and trended towards a significant positive association with IFNy+IL17+ CD4+ T cells (i.e. inflammatory Th17 cells) (Fig. S8). L1FLnI_5q32o (Fig. S9) and LTR19_12p13.31 (Fig. S10) demonstrated no significant correlations with analyzed CD4+ T cell subset abundances in the gut. Furthermore, systemic markers of microbial translocation and inflammation including as plasma lipopolysaccharide (LPS) and interleukin (IL)-6, were analyzed and demonstrated insignificant associations with LTR19_12p13.31, L1FLnI_1q23.1s, or L1FLnI_5q32o expression (Fig. S11). Collectively, LTR19_12p13.31 and L1FLnI_1q23.1s, but not L1FLnI_5q32o, show significant associations with the various innate and adaptive immune cell frequencies in the gut of PLWH.

**Figure 3.**
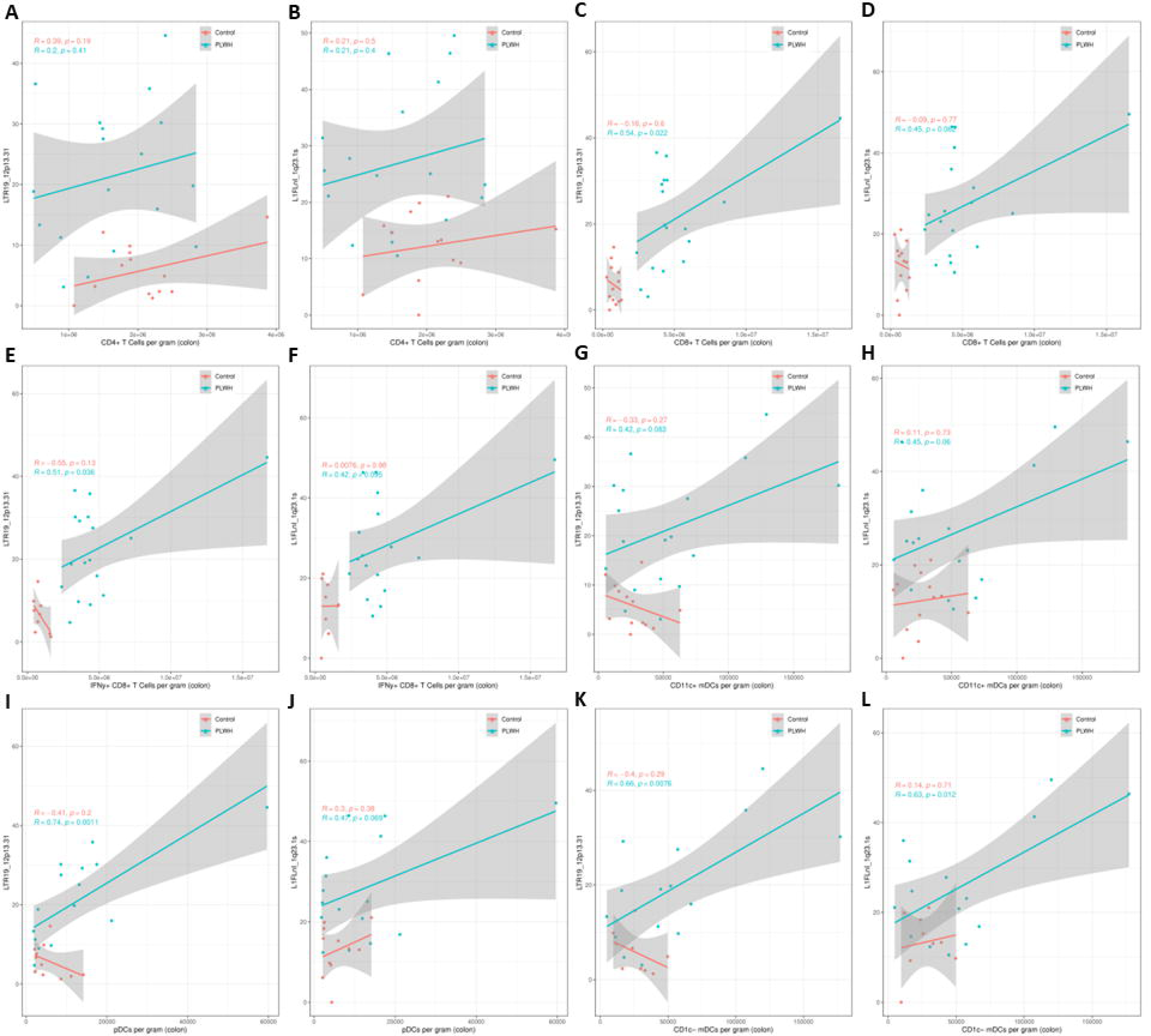
Correlation Between LTR19_12p13.31 and L1FLnI_1q23.1s expression and immune signatures of the gut microenvironment in people living with HIV-1. For all samples uninfected controls (red) (n = 13) vs PLWH (blue) (n = 19). Scatterplot demonstrating the CD4+ T Cells per gram on the X axis and normalized expression of LTR19_12p13.31 in colon biopsies on the Y axis (A). Scatterplot demonstrating the CD4+ T Cells per gram on the X axis and normalized expression of L1FLnI_1q23.1s in colon biopsies on the Y axis (B). Scatterplot demonstrating the CD8+ T Cells per gram on the X axis and normalized expression of LTR19_12p13.31 in colon biopsies on the Y axis (C). Scatterplot demonstrating the CD8+ T Cells per gram on the X axis and normalized expression of L1FLnI_1q23.1s in colon biopsies on the Y axis (D). Scatterplot demonstrating the IFNy+CD8+T Cells per gram on the X axis and normalized expression of LTR19_12p13.31 in colon biopsies on the Y axis (E). Scatterplot demonstrating the IFNy+CD8+ T Cells per gram on the X axis and normalized expression of L1FLnI_1q23.1s in colon biopsies on the Y axis (F). Scatterplot demonstrating the CD11c+ mDCs per gram on the X axis and normalized expression of LTR19_12p13.31 in colon biopsies on the Y axis (G). Scatterplot demonstrating the CD11c+ mDCs per gram on the X axis and normalized expression of L1FLnI_1q23.1s in colon biopsies on the Y axis (H). Scatterplot demonstrating the pDCs per gram on the X axis and normalized expression of LTR19_12p13.31 in colon biopsies on the Y axis (I). Scatterplot demonstrating the pDCs per gram on the X axis and normalized expression of L1FLnI_1q23.1s in colon biopsies on the Y axis (J). Scatterplot demonstrating the CD1c-mDCs per gram on the X axis and normalized expression of LTR19_12p13.31 in colon biopsies on the Y axis (K). Scatterplot demonstrating the CD1c-mDCs per gram on the X axis and normalized expression of L1FLnI_1q23.1s in colon biopsies on the Y axis (L).

## Discussion

In this study, we leverage modulations in ERE activity at the RNA level in the GI tract of untreated PLWH with frequencies of immune cell populations that are likely critical players in driving HIV-1 gut pathogenesis^11^. As the derepression of EREs may possess immunomodulatory roles in human diseases^42,61^, our results suggest further investigation into their activity in HIV-1 pathogenesis. By analysis of RNA sequencing from colon biopsies of uninfected controls vs untreated PLWH, we identified 59 differentially expressed EREs associated with HIV. By finding correlations between their colonic expression and circulating viral load, we were able to narrow further investigations to focus on 3 EREs upregulated in PLWH that might indicate the severity of HIV-1 pathogenesis in the gut, or function as markers of immune deregulation. The EREs LTR19_12p13.31 and L1FLnI_1q23.1s best correlated with cellular abundances including higher frequencies of pDCs, CTLs, and CD1c-mDCs. L1FLnI_5q32o demonstrated no associations with these cellular markers, however, did show associations with differential abundances of CD4+ T cell Th subsets that were not found with LTR19_12p13.31 and L1FLnI_1q23.1s expression. By considering their expression in PBMCs from the same study participants, we observed a significant upregulation in L1FLnI_1q23.1s expression and a non-significant upregulation in LTR19_12p13.31. In accordance with previous studies^22^, we observe significant alterations in ERE activity at the RNA level in the PBMC fraction of PLWH and uninfected controls (Fig. S12). This suggests future use of L1FLnI_1q23.1s as a potential circulating biomarker for deregulations of immunity in the gut microenvironment of PLWH, while LTR19_12p13.31 is more likely an intrinsic marker specific to the GI tract. Collectively, we find significant associations between LTR19_12p13.31 and L1FLnI_1q23.1s and cellular dynamics of the GALT in PLWH, further emphasizing their identity as biomarkers or potential mediators of immune deregulation.

Previous studies have demonstrated that ERE activity is widely changed in response to the HIV-1 lifecycle^21,22,27,29,31^ and certain HIV-1 associated comorbidities^62,63^. ERE expression is also susceptible to changes the in epigenetics and immunity that are elicited during viral infections and the host response^64^. While it remains unknown if the changes in expression for most EREs result from a bystander effect or by a cooption of host immunity in HIV-1 infection, their activity can be incorporated to better characterize poorly understood immunopathologies in viral infections. Here, we demonstrate that significant correlations can be established between differentially expressed EREs and cellular markers that likely contribute to GI immunopathologies of untreated HIV-1 infection. Some limitations of this study concern bulk RNA sequencing approaches utilized, leaving the data unable to discern what cell types are responsible for the overexpression of differentially expressed EREs. As the cellular composition of the sequenced tissues fluctuates with disease severity, it is unclear whether the phenotype or mere abundance of disproportionate cell types, such as pDCs and CTLs, is responsible for their expression. Future studies that accurately quantify ERE expression from single cell RNA sequencing data that were not available from this cohort would provide further insight into the activation state of these EREs in the GI tract of PLWH. Additionally, due to the poorly defined molecular characteristics and low copy number of LTR19 HERV sequences, exceedingly little is known about their potential roles in health and disease. LTR19 sequences are a member of the HERVFA clade and are estimated to only possess roughly ∼15 copies in the human genome^65^.

In conclusion, this study provides the first correlative link between ERE expression and the deregulations of GALT observed in untreated HIV-1 infection. While EREs can be modulated in context dependent manners by various aspects of HIV-1 infection, their potential roles in systemic immunity in the GI tract have remained undetermined. We also demonstrate that the IFN-I response mediates ERE expression in uninfected gut CD4+ T cells, however this only appears to be responsible for a distinct subset of roughly 7 differentially expressed elements. It is thus unclear if the HIV-1 lifecycle or IFN-I activity on CD4-cell types in the GALT are responsible for changes in ERE activity. We find that LTR19_12p13.31 and L1FLnI_1q23.1s are promising markers of HIV-1 pathogenesis in the gut and may possess undetermined roles in determining cellular composition. This study also provides one of the first observations of a differentially expressed LTR19 elements in a human disease, thus emphasizing the importance of their further study.

## Contributors

ND co-designed the study, co-prepared the first manuscript draft, co-performed data analyses, co-curated code, led statistical analyses, and curated figures. TF co-prepared the first manuscript draft, co-performed data analyses, co-performed statistical analysis, co-curated code, and provided intellectual input critical to analysis. SM co-designed the study, co-prepared the first manuscript draft, co-performed data analyses, and provided intellectual input critical to data interpretation. NL co-performed analyses and co-curated code. KG provided intellectual input critical to hypothesis generation and co-provided data access. KLM provided intellectual input critical to hypothesis generation and co-provided data access. BSB provided intellectual input critical to hypothesis generation and co-provided data access. MLB provided intellectual input critical to hypothesis testing. SMD provided intellectual input critical to hypothesis generation and testing, co-provided data access, and performed immunological analysis of the gut microenvironment. CCW provided intellectual input critical to hypothesis generation and co-provided data access. DFN co-designed the study and co-prepared the first manuscript draft. MLS co-designed the study, co-prepared the first manuscript draft, and co-provided data access. All authors contributed to data interpretation, discussion regarding significances, and editing of the manuscript. All authors have read and approved the final manuscript.

## Data Availability

All data produced in the present study are available upon reasonable request to the authors

## Acknowledgments

These works are supported by US NIH grants NCI CA260691 (DFN) and NIAID UM1AI164559 (DFN).

## Data Sharing Statement

FASTQ from the bulk RNA sequencing reanalyzed by this study can be accessed at the NCBI sequence read archive (SRA) under PRJNA558974 and PRJNA558500. Code for analysis can be accessed at https://github.com/NicholasDopkins/JuneHIVMucosa. Details on running the Telescope pipeline can be accessed at https://github.com/mlbendall/telescope.

## Supplemental Legends

**Supplemental Table 1.**
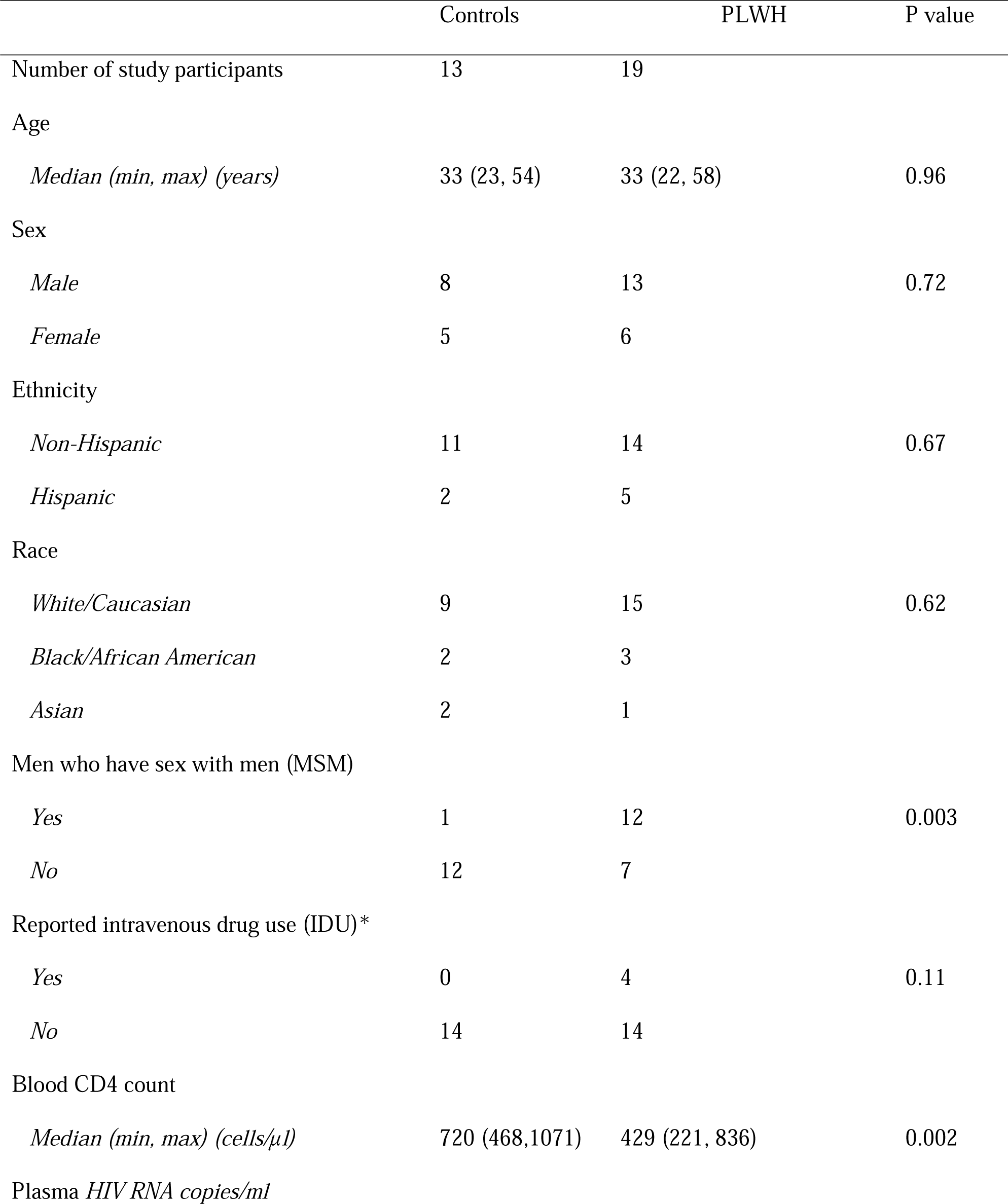
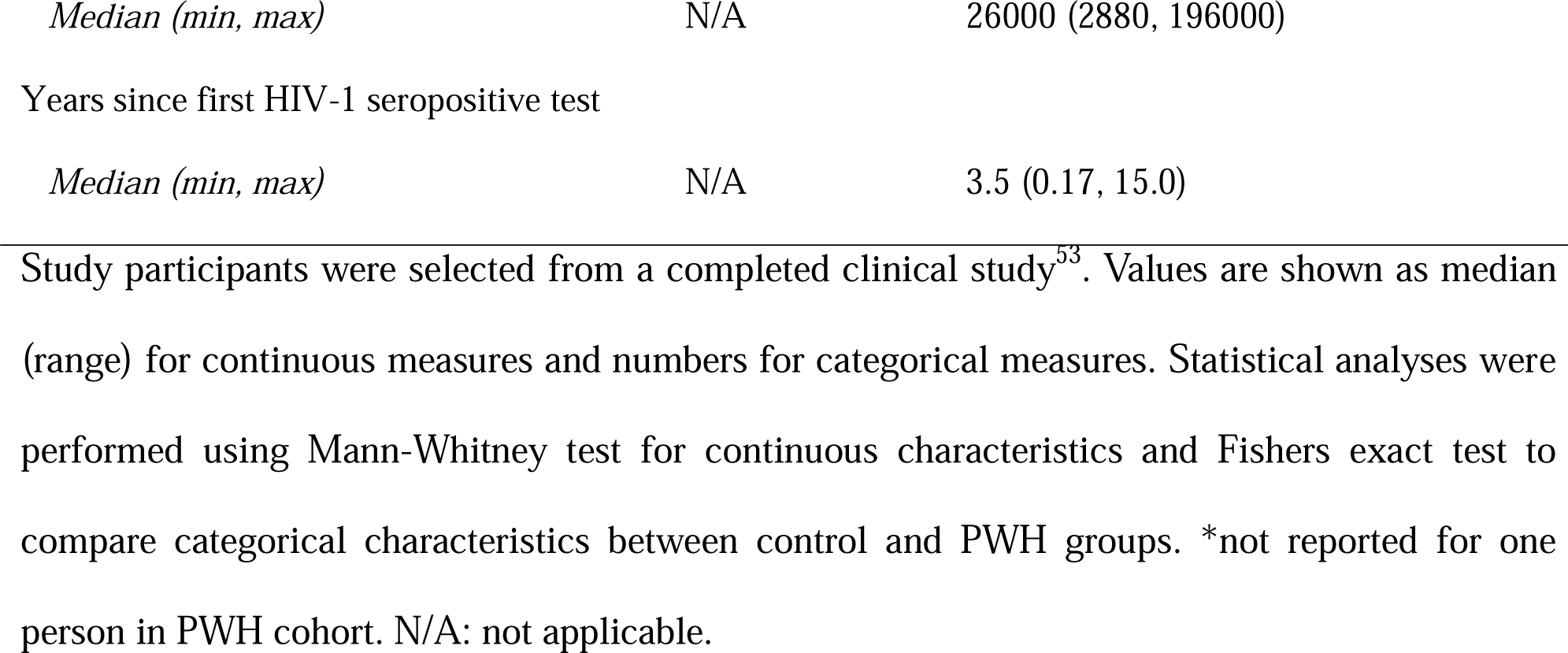

|  | Controls | PLWH | P value |
| --- | --- | --- | --- |
| Number of study participants | 13 | 19 |  |
| Age |  |  |  |
| *Median (min, max) (years)* | 33 (23, 54) | 33 (22, 58) | 0.96 |
| Sex |  |  |  |
| *Male* | 8 | 13 | 0.72 |
| *Female* | 5 | 6 |  |
| Ethnicity |  |  |  |
| *Non-Hispanic* | 11 | 14 | 0.67 |
| *Hispanic* | 2 | 5 |  |
| Race |  |  |  |
| *White/Caucasian* | 9 | 15 | 0.62 |
| *Black/African American* | 2 | 3 |  |
| *Asian* | 2 | 1 |  |
| Men who have sex with men (MSM) |  |  |  |
| *Yes* | 1 | 12 | 0.003 |
| *No* | 12 | 7 |  |
| Reported intravenous drug use (IDU)* |  |  |  |
| *Yes* | 0 | 4 | 0.11 |
| *No* | 14 | 14 |  |
| Blood CD4 count |  |  |  |
| *Median (min, max) (cells/μl)* | 720 (468,1071) | 429 (221, 836) | 0.002 |
| Plasma *HIV RNA copies/ml* |  |  |  |

|  |  |  |
| --- | --- | --- |
| *Median (min, max)* | N/A | 26000 (2880, 196000) |

Years since first HIV-1 seropositive test
|  |  |  |
| --- | --- | --- |
| *Median (min, max)* | N/A | 3.5 (0.17, 15.0) |

Study participants were selected from a completed clinical study<sup>53</sup>. Values are shown as median (range) for continuous measures and numbers for categorical measures. Statistical analyses were performed using Mann-Whitney test for continuous characteristics and Fishers exact test to compare categorical characteristics between control and PWH groups. \*not reported for one person in PWH cohort. N/A: not applicable.

## Research in Context

### Evidence before this study

Previous studies have demonstrated that HIV-1 infection alters the physiology of the GI tissues, where it forms quiescent reservoirs. In chronic HIV-1, the sustainment of antiviral immune responses through poorly understood mechanisms likely contributes to pathogenesis and prohibits effective clearance of the viral reservoirs. EREs comprise a substantial portion of the human genome that has been implicated in HIV-1 pathogenesis but remains understudied in comparison to coding genes. Therefore, studying the activity of EREs in tissues such as the GI tract may help further define the physiology of immunopathologies that contribute to HIV-1 reservoir persistence.

### Added value of this study

Colon pinch biopsies from PLWH demonstrate differential expression of 59 EREs when compared to uninfected controls. Of these 59, 3 possess near-significant correlations with viral load. Further analysis into cellular composition of the gut microenvironment demonstrates that 2 the EREs LTR19_12p13.31 and L1FLnI_1q23.1s possess significant associations with immune cell phenotypes indicative of localized HIV-1 pathogenesis. Therefore, high expression of LTR19_12p13.31 and L1FLnI_1q23.1s can function as markers of GI HIV-1 reservoirs, as well as potentially possess undefined roles in human health.

### Implications of all the available evidence

Recurrent observations suggest pathophysiological roles for differentially expressed EREs in diseased tissues. In HIV-1 infection, EREs can be over expressed due to trans-activation of HERV LTRs and shifts in the regulome that permit the expression of typically silent elements. However, it has not been established how EREs may be deregulated in the gut where immunopathologies persist surrounding viral reservoirs. Correlations between ERE expression and cellular composition demonstrate that certain EREs may be indicative of cell type abundances thought to drive GI-specific immunopathologies resultant from HIV-1 infection.

