## Supplemental Figures for "Endogenous Retroelement Expression in the Gut Microenvironment of People Living with HIV-1"

### Slide 1
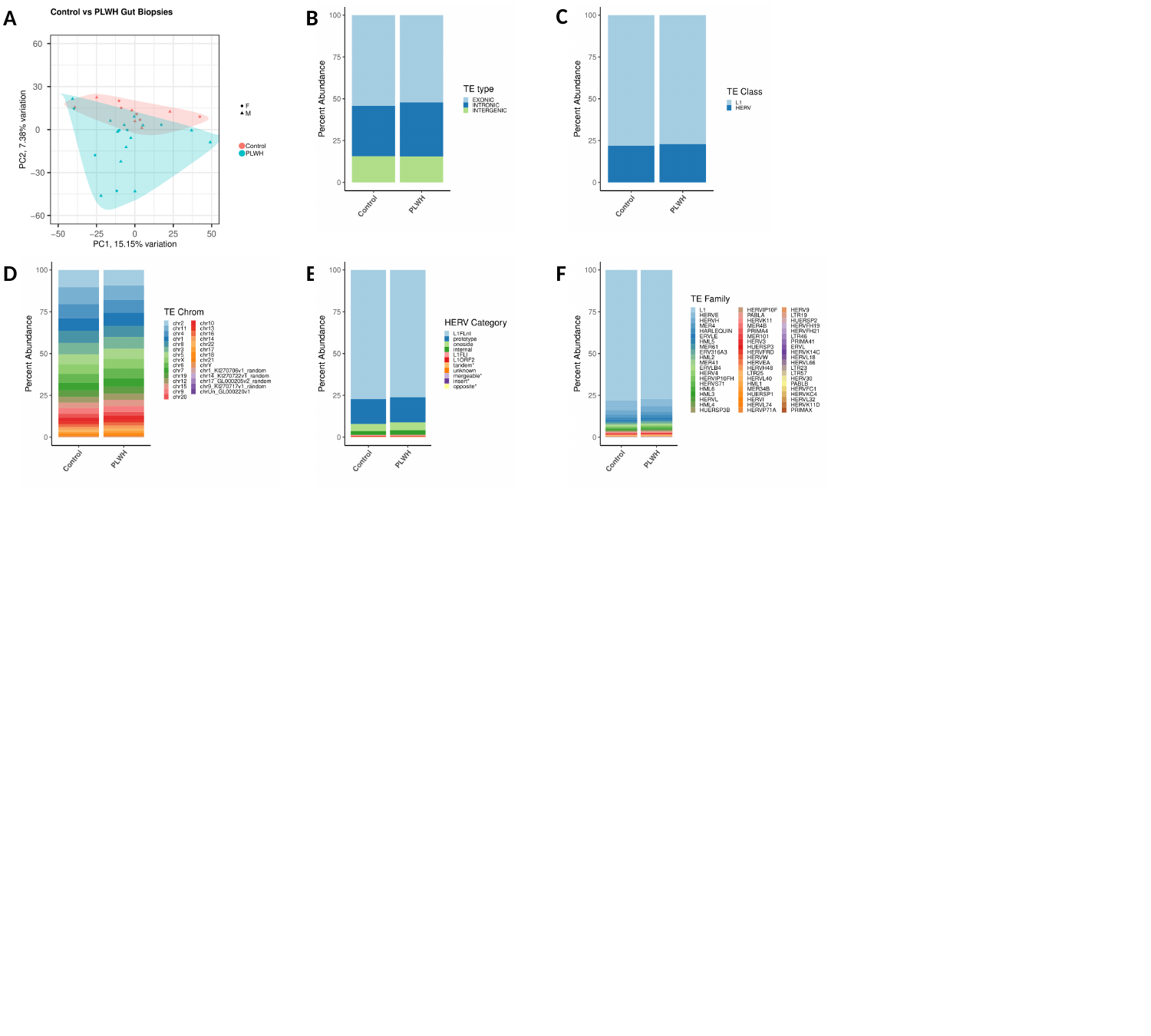

C
A
B
D
E
F

### Slide 2
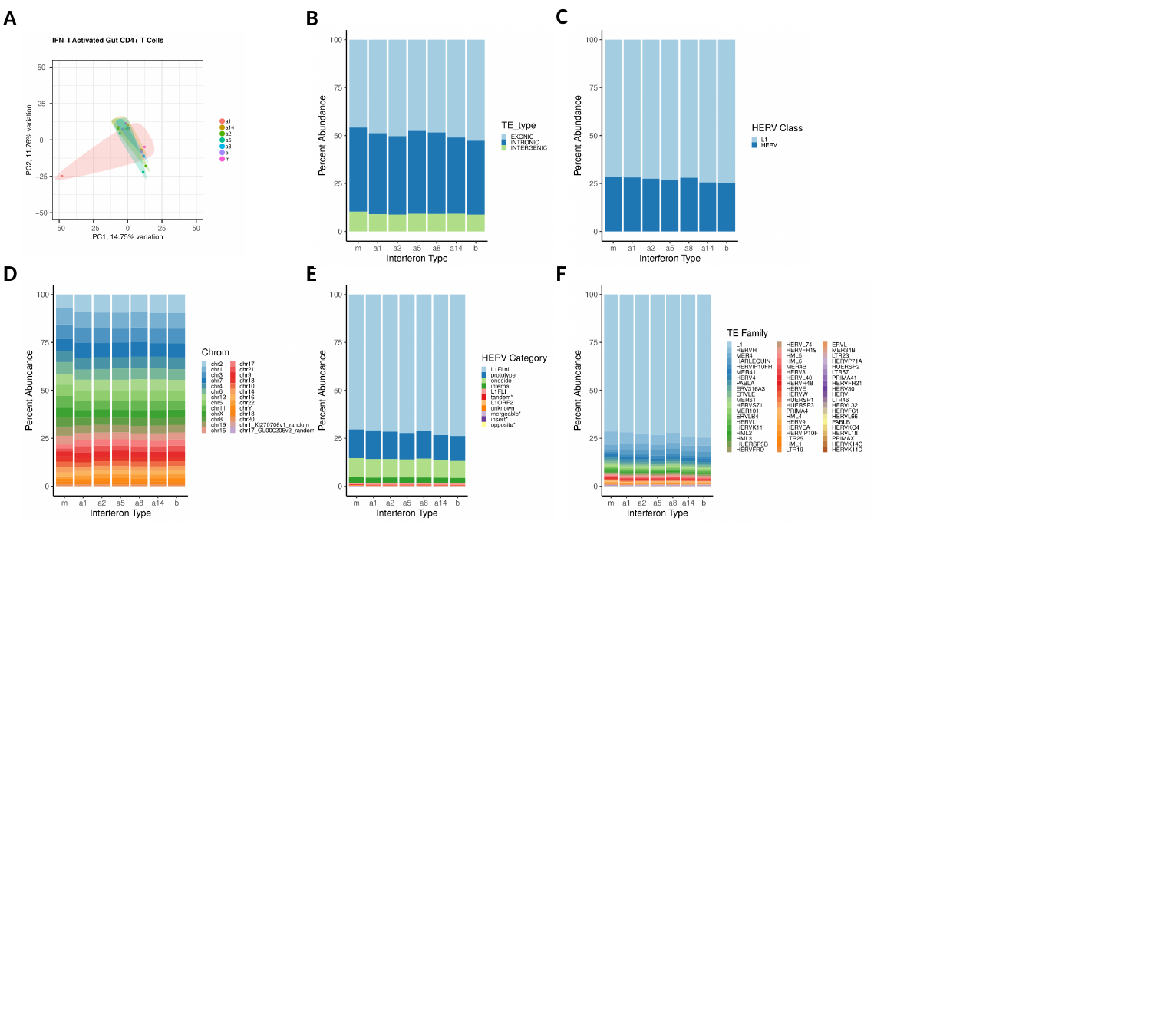

C
A
B
D
E
F

### Slide 3
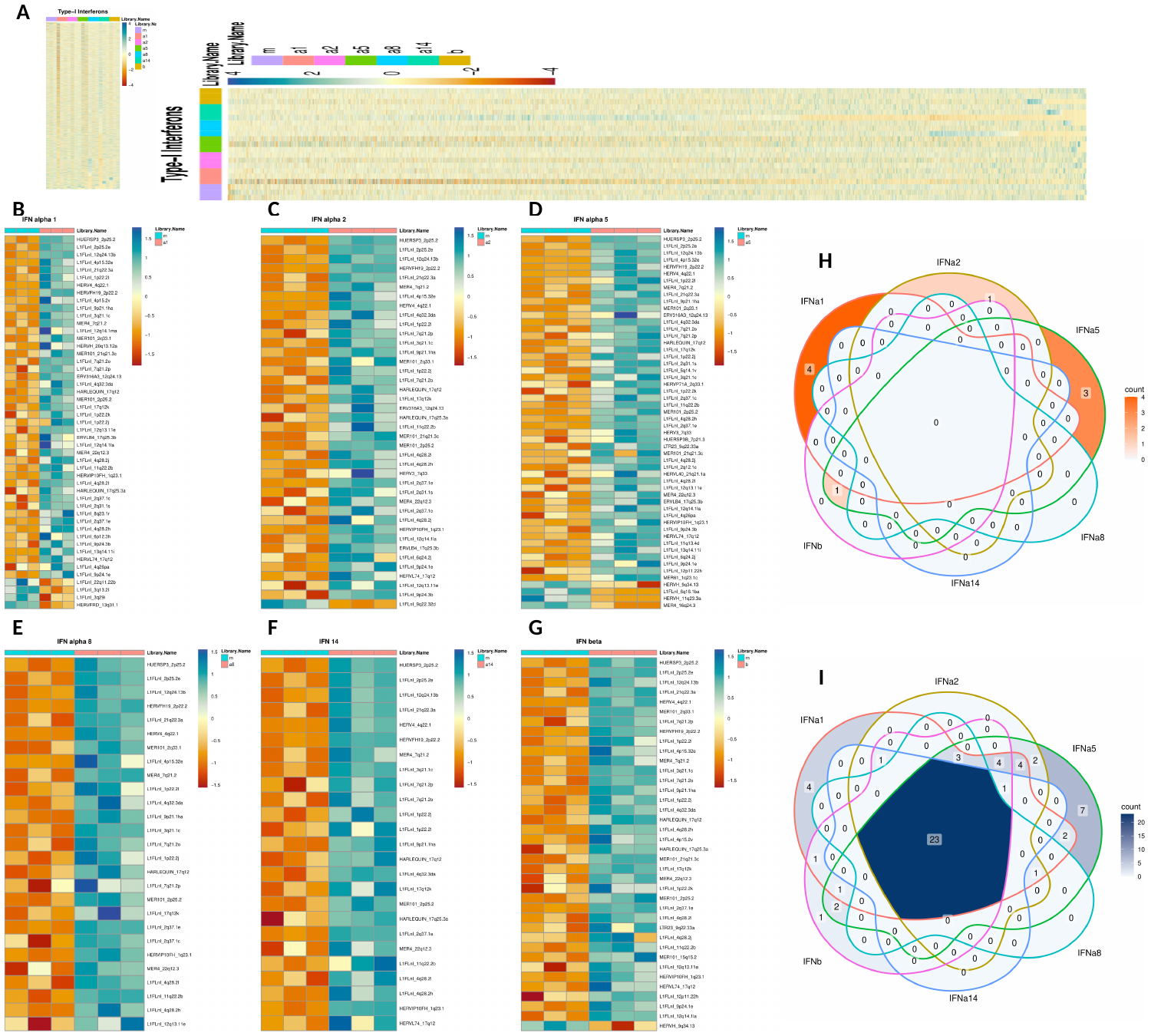

A
B
C
D
H
E
F
G
I

### Slide 4
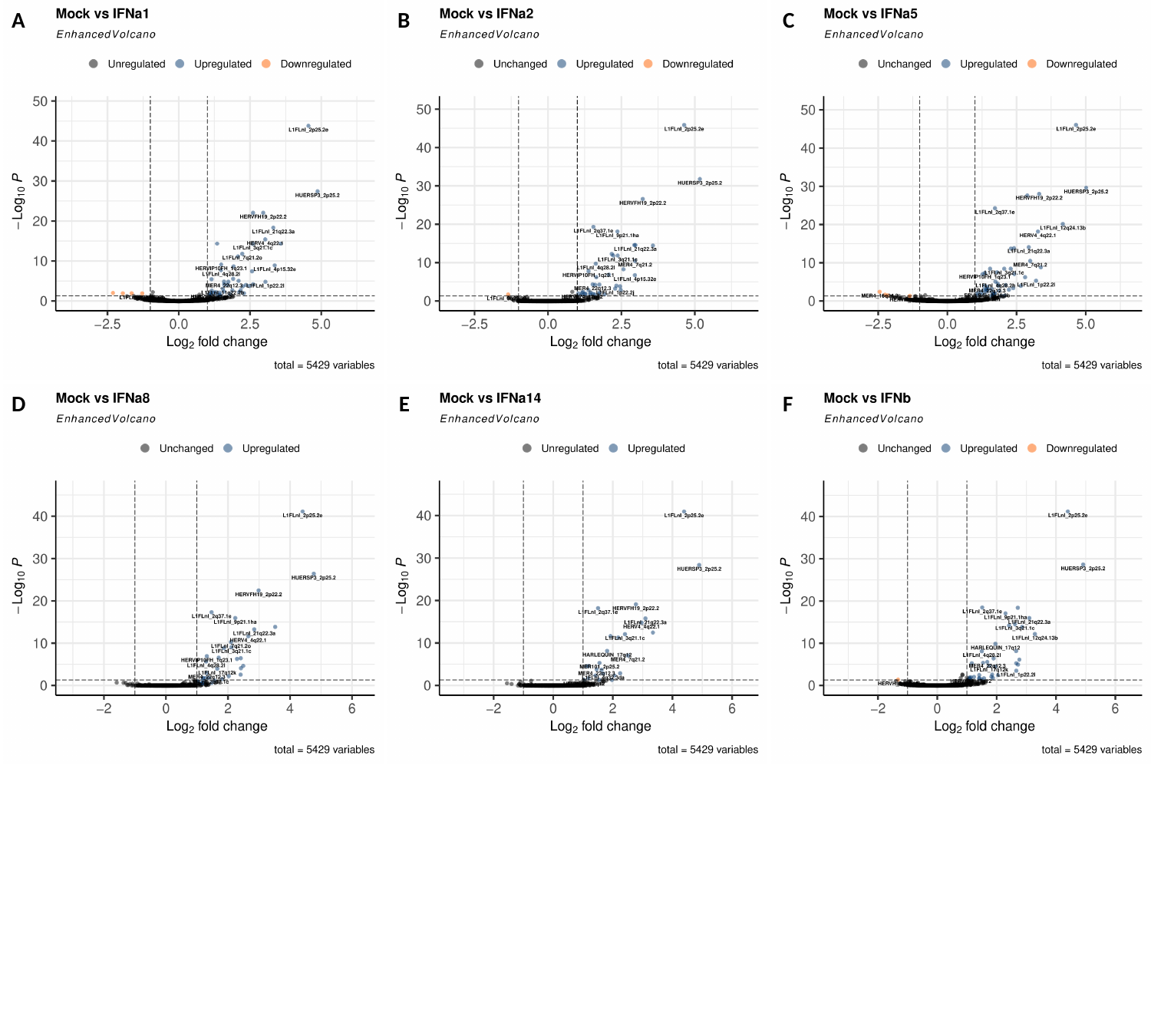

A
B
C
D
E
F

### Slide 5
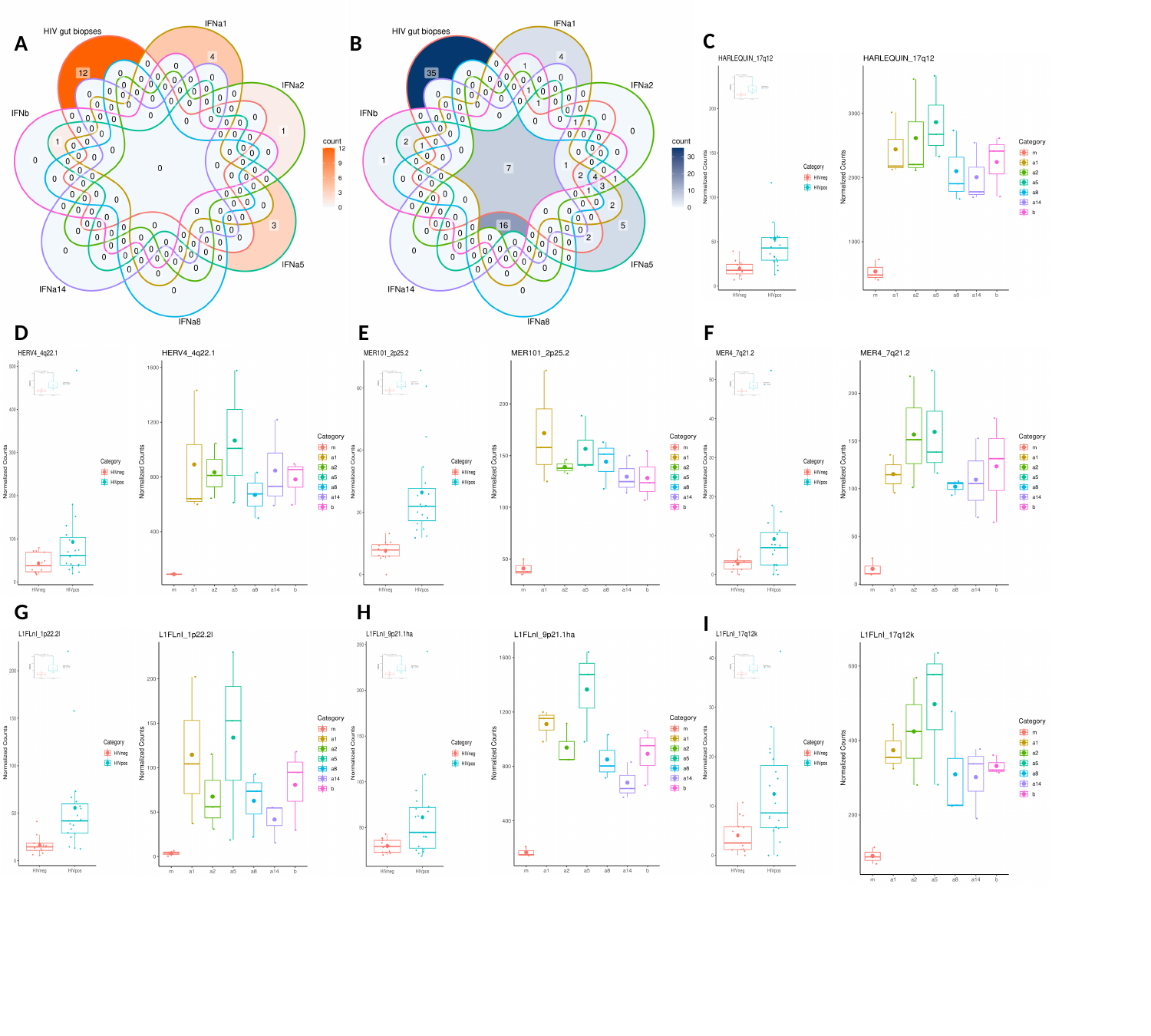

C
A
B
D
E
F
G
H
I

### Slide 6
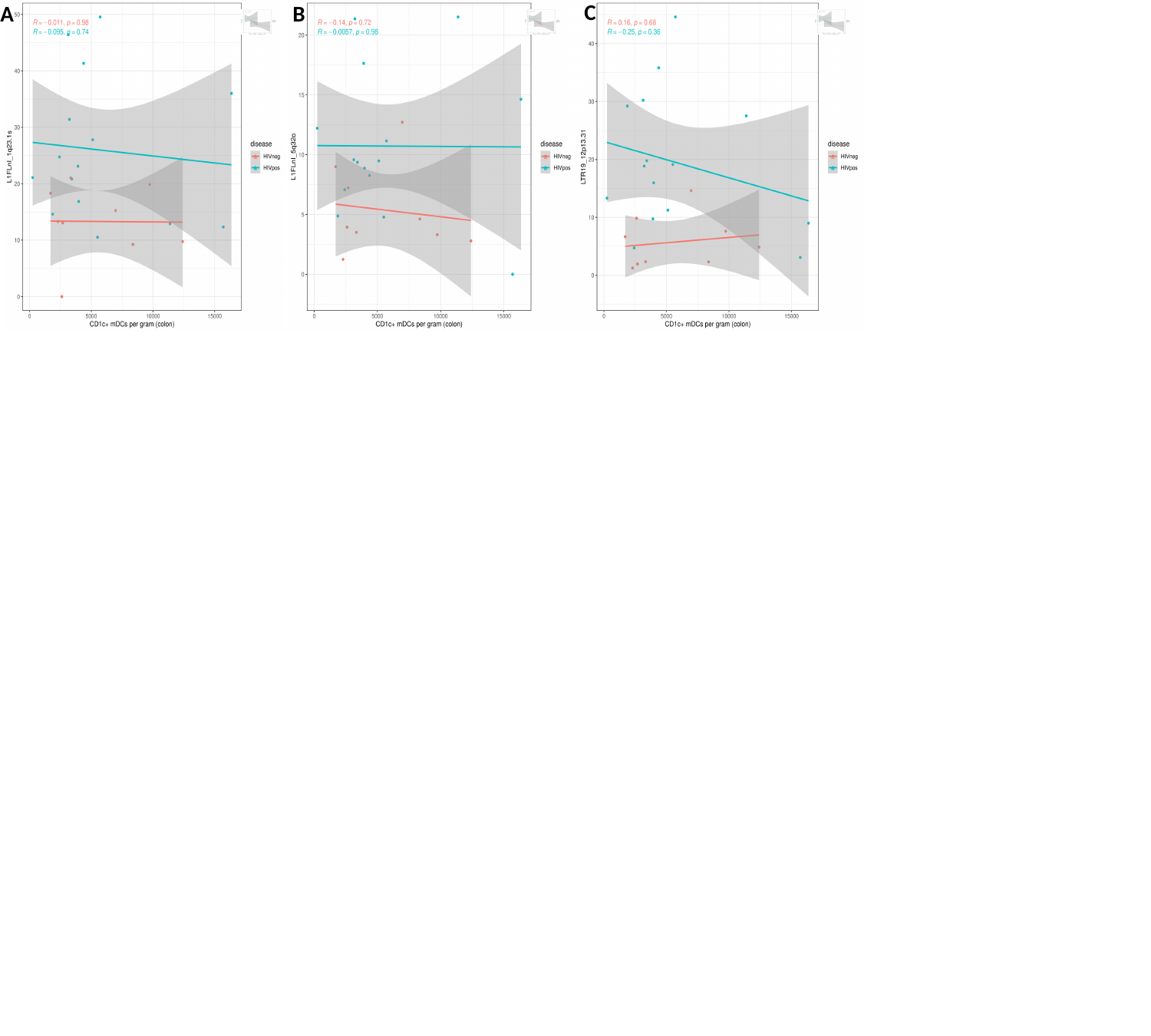

C
A
B

### Slide 7
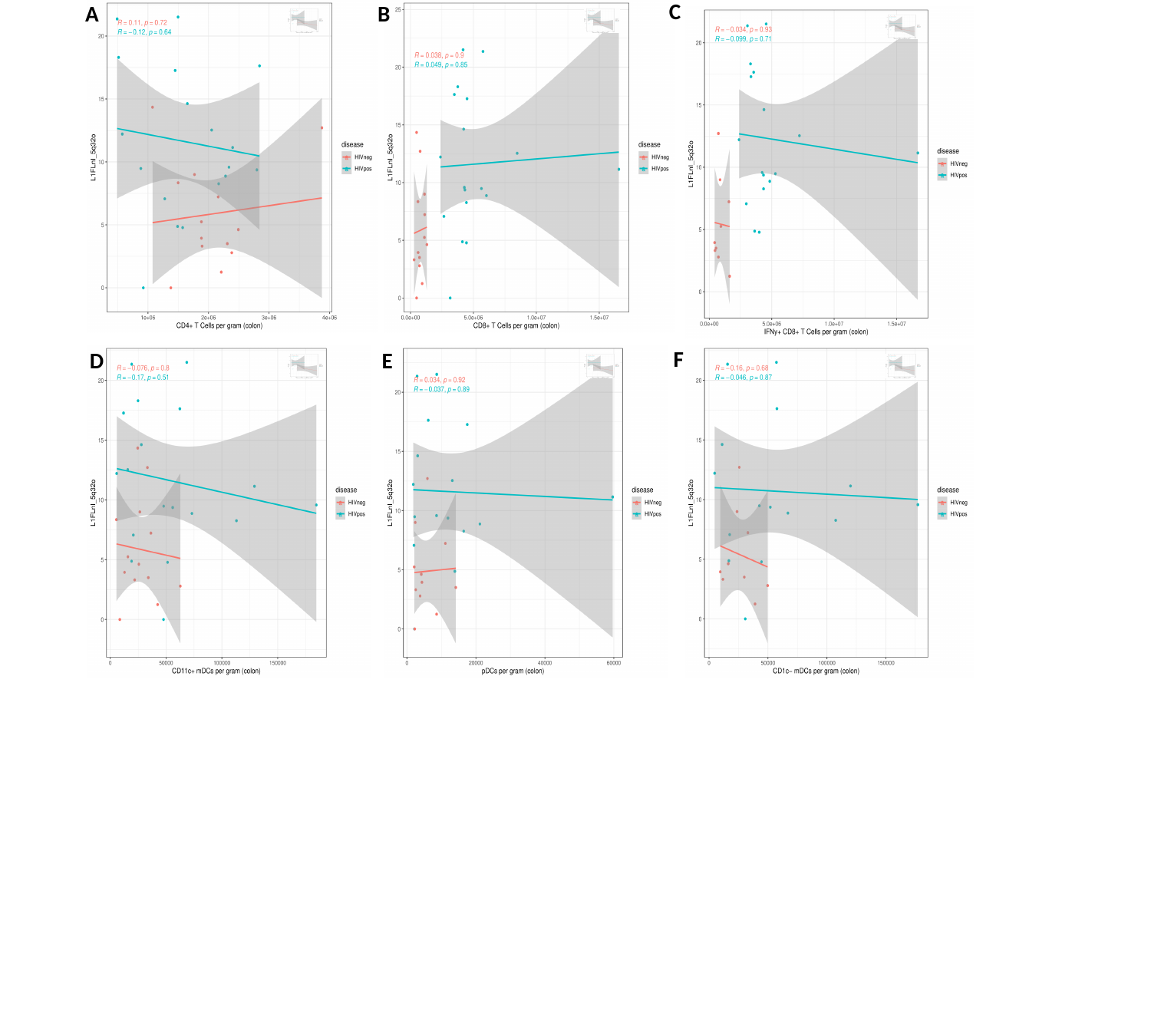

C
A
B
F
D
E

### Slide 8
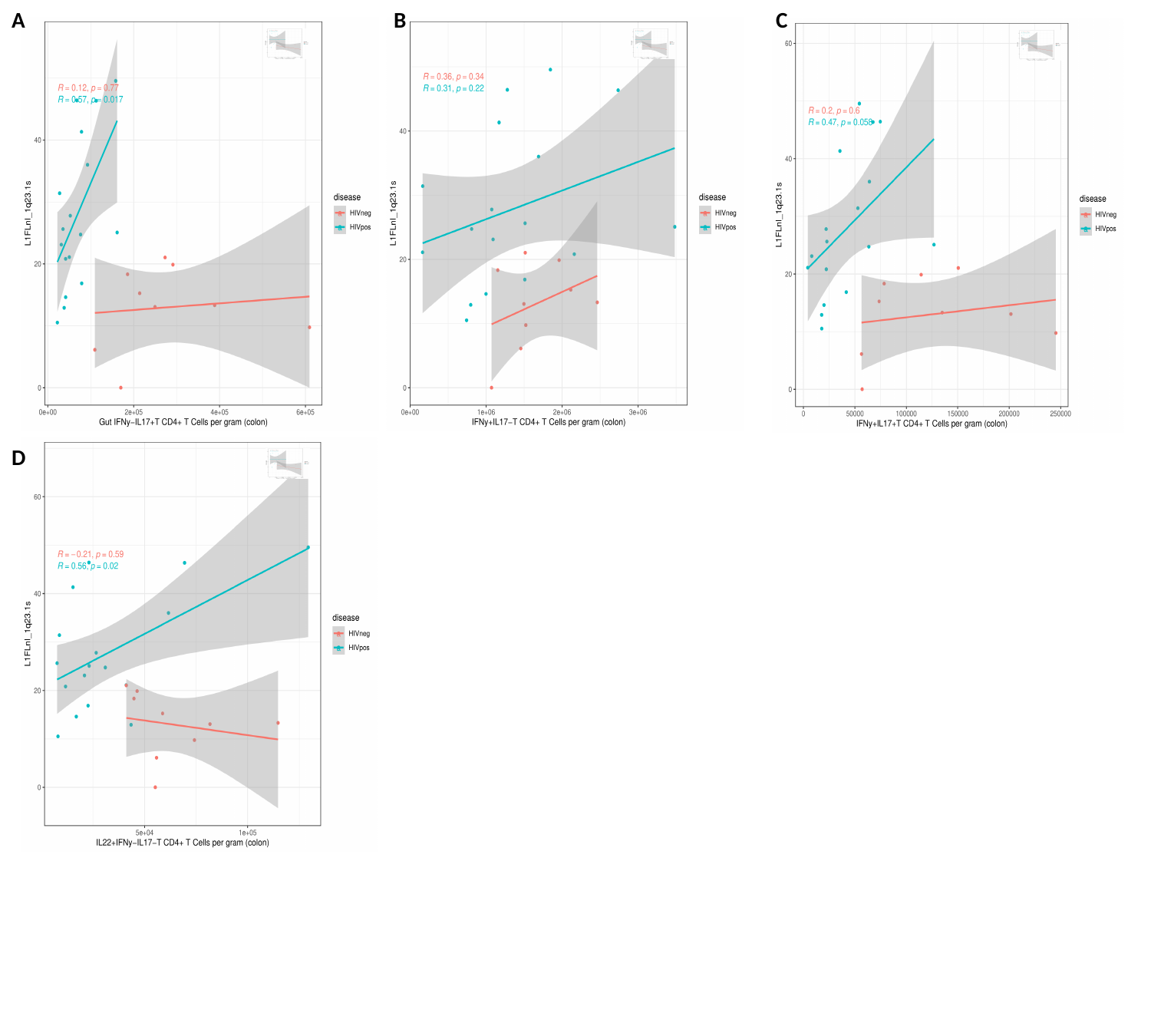

A
B
C
D

### Slide 9
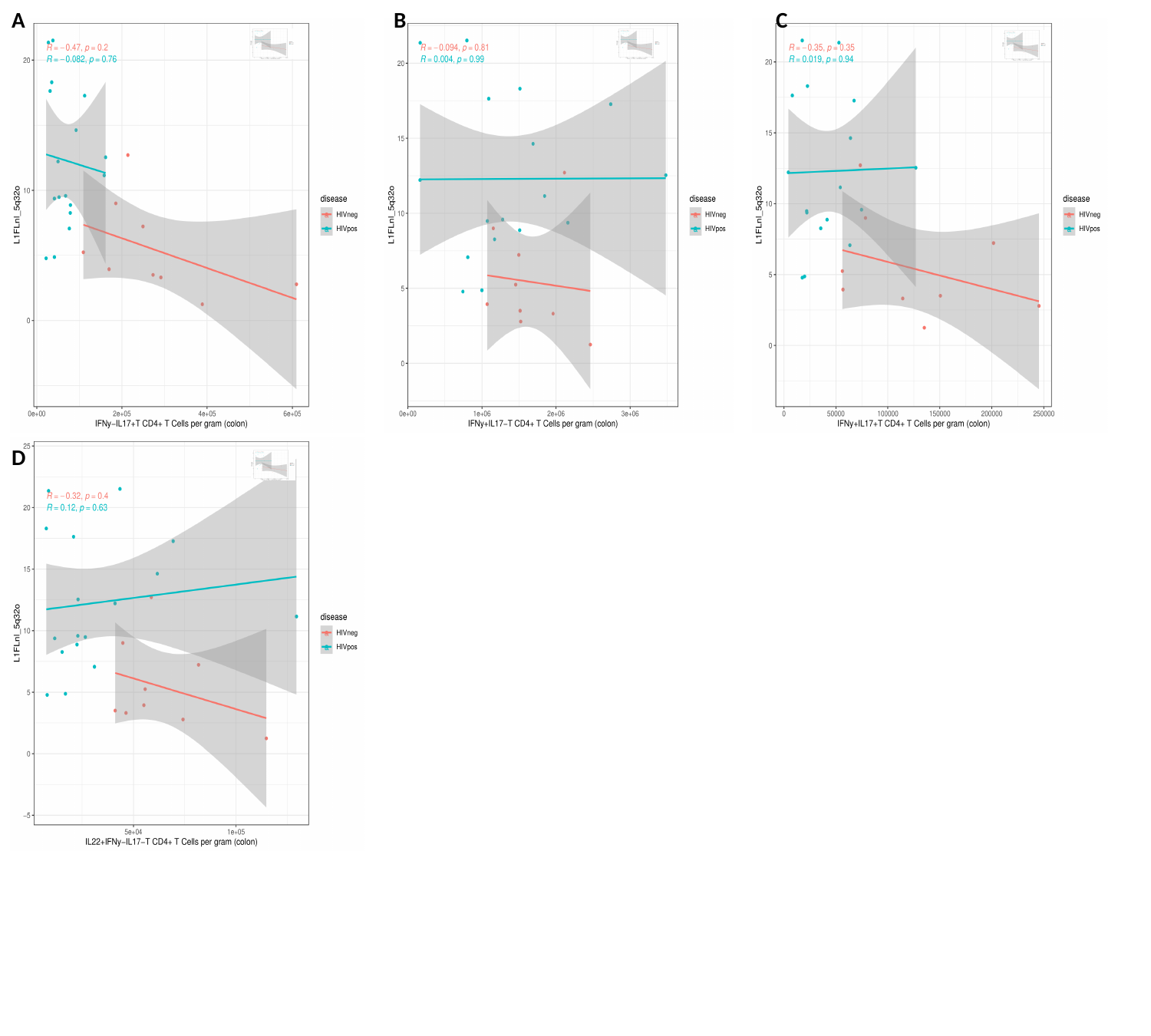

A
B
C
D

### Slide 10
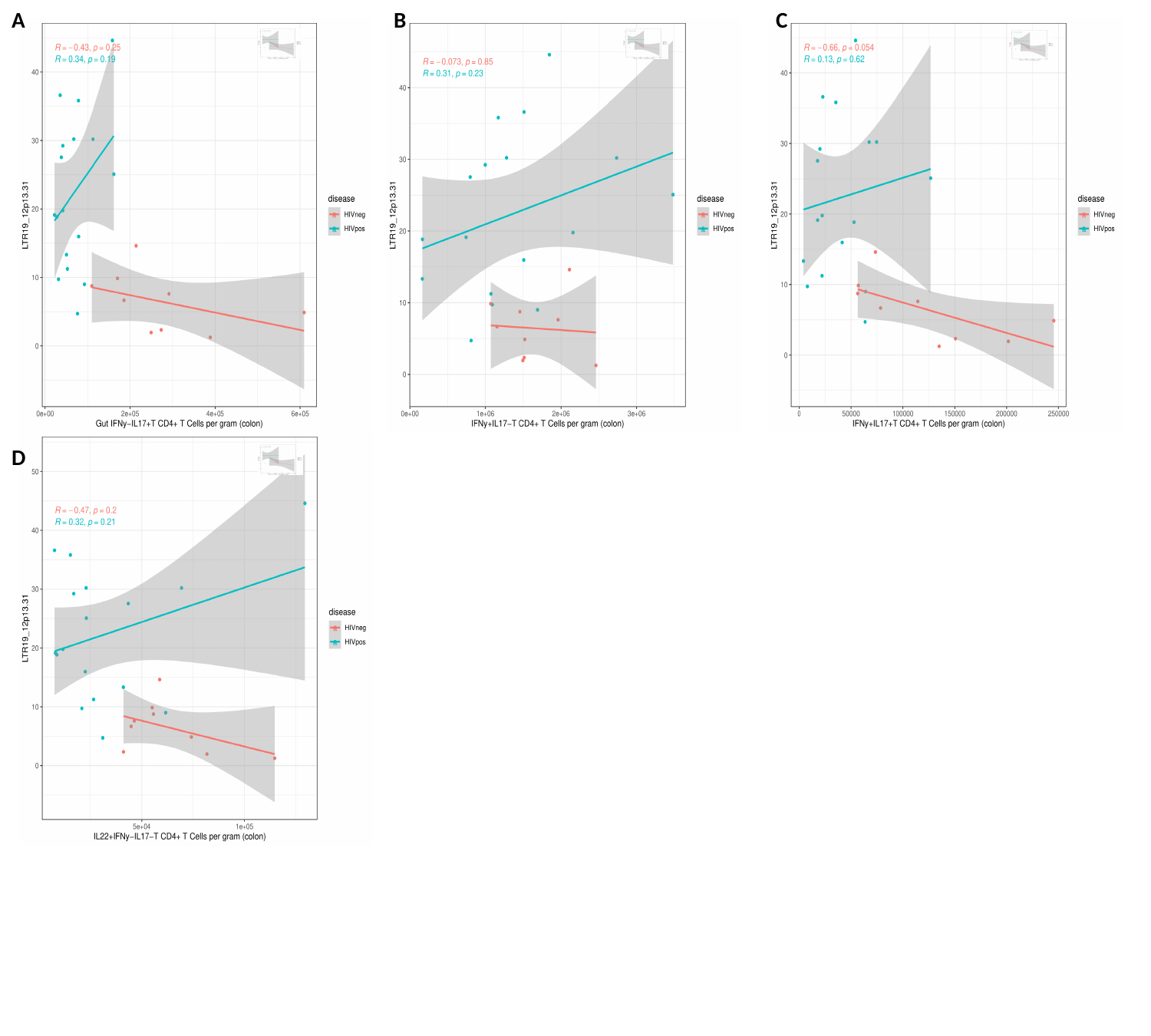

A
B
C
D

### Slide 11
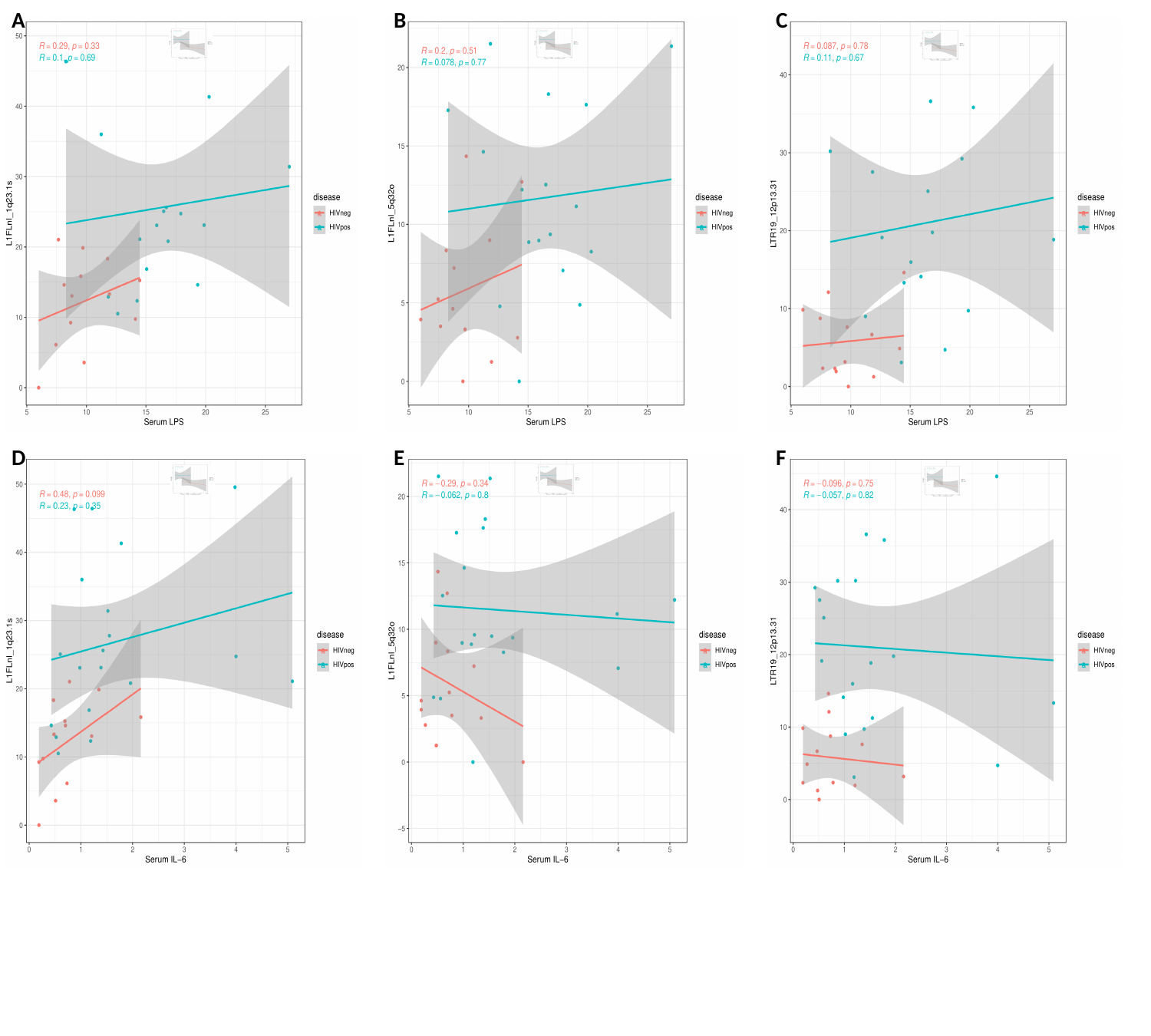

A
B
C
D
E
F

### Slide 12
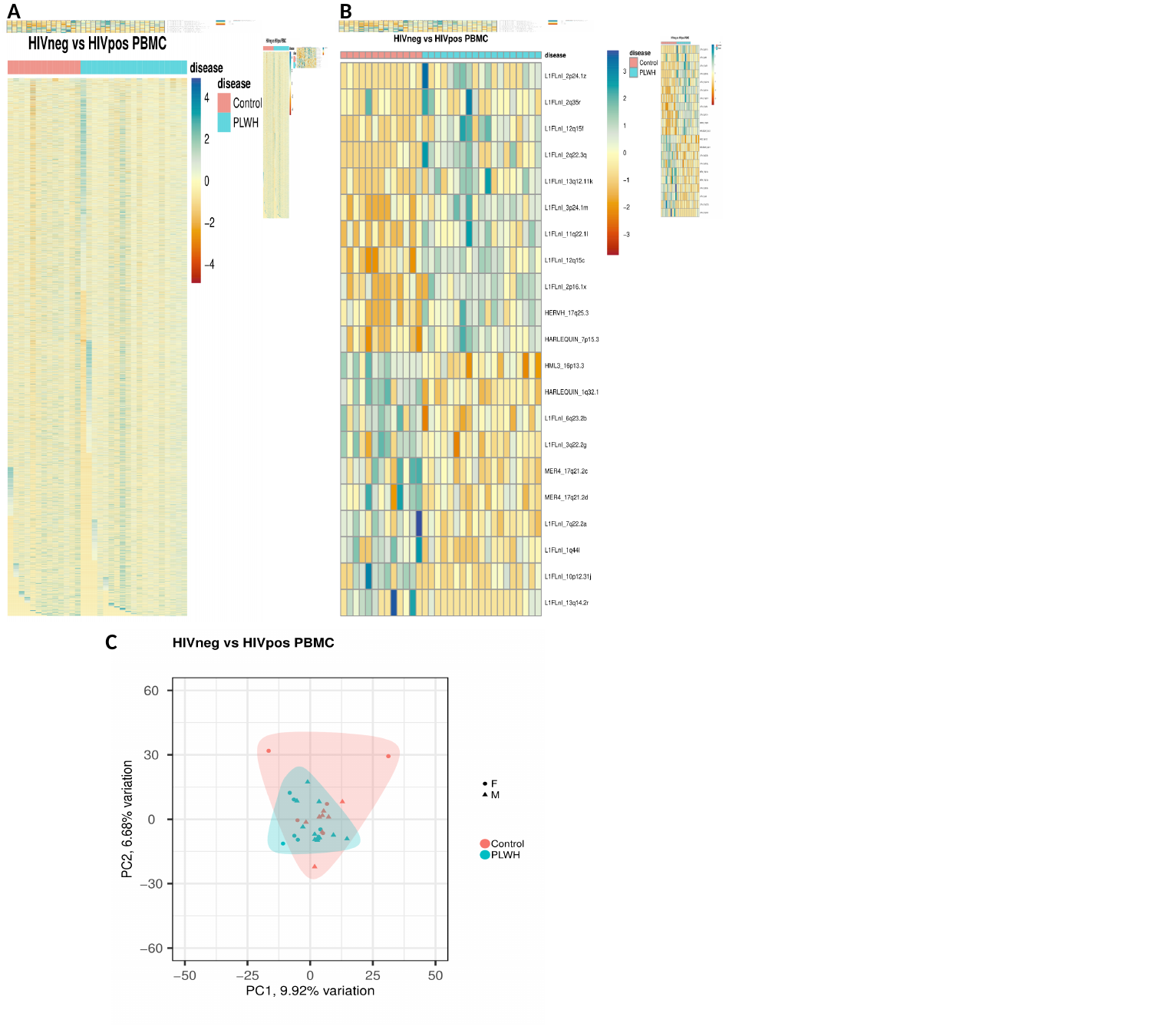

A
B
C
